# Intimate partner violence in Nepal: Analysis of Nepal Demographic and Health survey 2022

**DOI:** 10.1101/2024.01.09.24301029

**Authors:** Parash Mani Sapkota, Achyut Raj Pandey, Bikram Adhikari, Grishu Shrestha, Reecha Piya, Bipul Lamichhane, Shristi Garu, Deepak Joshi, Sushil Chandra Baral

## Abstract

**Introduction:** Intimate partner violence (IPV) is a major public health issue in Nepal. IPV has social and economic impacts among women, adolescent girls, wider society. In this study we aimed to determine factors associated with intimate partner violence among women aged 15-49 currently in intimate relationship.

**Methods:** We conducted secondary data analysis of the Nepal Demographic and Health Survey 2022. IPV was measured in three domains-experience of physical violence, emotional violence, and sexual violence. Weighted univariate and multivariate logistic regression analysis was applied to determine factors associated with intimate partner violence. The results of logistic regression were presented as odds ratio and their 95% CI.

**Results:** Of 3853 women, 27.2% had experienced any form of IPV. The prevalence of physical violence, emotional violence and sexual violence were 23.2%, 12.8 and 7.1% respectively. Higher odds of physical violence were reported among women aged 35-49 years (AOR: 2.13, 95%CI: 1.58-2.87), women without formal education (AOR: 1.51, 95%CI: 1.10-2.06) and women who justified wife beating (AOR: 1.23, 95%CI: 1.00-1.52). Women from poor households (AOR: 1.61, 95%CI: 1.12-2.35) and women with uneducated partners (AOR: 1.66, 95%CI: 1.08-2.58) were at higher risk of experiencing sexual violence. Women with unemployed husbands reported higher risk of physical violence (AOR: 2.72, 95%CI: 1.45-5.06) and emotional violence (AOR: 1.61, 95%CI: 1.12-2.35). Women who were afraid of their partners, women who experienced parental violence, women with partners who were sometimes or often drunk, and women with controlling partners were many folds more likely to experience all forms of IPV.

**Conclusion:** The prevalence of all forms of IPV were high among Nepalese women. Various sociodemographic, partner-related, and women empowerment related factors were positively associated with experiencing IPV. Acknowledging and addressing all these factors is essential to mitigating the alarming rates of IPV in this population.

## Introduction

Violence against women and girls (VAWG) is one of the most widespread and persistent form of human right violations in our world today which remains largely unreported due to factors such as impunity, silence, stigma and shame [1]. The UN General Assembly issued the Declaration on the Elimination of Violence Against Women in 1993 defining it as “any act of gender-based violence that results in, or is likely to result in, physical, sexual or psychological harm or suffering to women, including threats, coercion or arbitrary deprivation of liberty, whether occurring in public or in private life” [2]. The 2030 UN Agenda for Sustainable Development Goals (SDGs), calls for the elimination of all forms of violence against women and girls in both the public and private spheres, including trafficking, sexual exploitation, or other forms of exploitation in target 5.2 under goal 5 [3]. The first indicator of the target (5.2.1) specifically focuses on IPV, requiring regular reporting on “the proportion of ever-partnered women and girls aged 15 years and above subjected to physical, sexual or psychological violence by a current or former intimate partner” [3]. The World Health Organization (WHO) defines Intimate Partner Violence (IPV) as behavior by an intimate partner or ex-partner causing physical, sexual, or psychological harm. This includes physical aggression, sexual coercion, psychological abuse and controlling behaviors [4].

Article 38 of the constitution of Nepal 2015 guarantees protection of women against physical, mental, sexual, psychological or any other form of violence or exploitation on any grounds and determine such acts to be punishable by law [5].

### Global Prevalence of IPV

Global Estimates of IPV perpetrated by men against women indicate that about one in three ever partnered women worldwide (30%) have experienced physical and/or sexual violence at some point in time by an intimate partner [6]. Lifetime estimates of IPV prevalence range from 20% in the Western Pacific, 22% in high-income nations and Europe, and 25% in the WHO Regions of the Americas to 33% in the WHO African region, 31% in the WHO Eastern Mediterranean region, and 33% in the WHO South-East Asia region. Lifetime estimates of IPV prevalence show regional variations with the rates being 20% in the Western Pacific Region, 22% in high-income nations and Europe, and 25% in the WHO Regions of the Americas. In contrast, the WHO African region has a rate of 33%, while the WHO Eastern Mediterranean region and the WHO South-East Asia region have rates of 31% and 33%, respectively [7]. An analysis of the WHO Global Database estimates that 27% of ever-partnered women aged 15-49 years have experienced physical or sexual IPV or both at least once in their lifetime [8]. The estimated prevalence of lifetime IPV and past year IPV in Southeast Asia region was 19% and 8% respectively [8].

### Prevalence of IPV in Nepal

According to the Nepal Demographic and Health Survey 2022, about 23% of women aged 15-49 years have experienced physical violence,7% have experienced sexual violence and 13% have experienced emotional violence in their lifetime [9]. Alarmingly, about 27% of the women have experienced some form of IPV in their lifetime. There was an increase of 3% in the percentage of ever-married women experienced IPV in the last 12 months from 14% in 2016 to 17% in 2022 [9]. Similarly, a study conducted among married women residing in Terai region of Nepal showed that almost 16% women reported physical violence, 18% reported sexual violence and 25% reported either or both violence perpetrated by their intimate partner [10].

## Methods

### Study Design

We performed secondary analysis of data from the Nepal Demographic and Health Survey (NDHS) 2022. NDHS 2022 is a nationally representative survey implemented by New ERA, a local research firm under the aegis of Ministry of Health and Population of Nepal. NDHS 2022 is the sixth comprehensive survey of its kind conducted as a part of Demographic and Health Survey (DHS) Program with technical support by ICF International and financial support by United States Agency for International Development (USAID) [9]. The dataset was publicly available on the ‘The DHS program’ website [11]. The details of the questionnaire and study methodology have been described in the study report [9].

### Sample size and sampling technique

In brief, a total of 14,845 women aged 15-49 were successfully interviewed, yielding a response rate of 97% and 95%, respectively. A total of 5,177 women were selected and interviewed for the domestic violence module out of total eligible 14,845 women aged 15-49. Only seven women were selected for the module but were not interviewed with the Woman’s Questionnaire, and six who were selected and interviewed with the Woman’s Questionnaire could not complete the module due to privacy concerns. Out of 5,177 women, 3853 women currently in an intimate relationship were exclusively selected due to the availability of information about their partner which allowed us to analyze the association of the partner characteristics with IPV. Partner information was captured only for women currently in an intimate relationship. Special weights were used to adjust for the selection of only one woman per household and to ensure the subsample was representative nationally.

### Data collection tool

The 2022 NDHS survey administered four major questionnaires: the Household’s Questionnaire, the Woman’s Questionnaire, the Man’s Questionnaire, and the Biomarker Questionnaire. The Woman’s Questionnaire was used to collect information on domestic violence from women aged 15-49 years.

The module on domestic violence was confined to respondents selected for the domestic violence module from the subsample of households selected for the men’s survey such that only one eligible woman per household was selected. The module was implemented only when privacy could be maintained, and the information collection process followed the WHO recommended guidelines and ethical standard [12].

More recently, the questionnaire module used to capture IPV in a DHS survey was revised to also capture IPV experienced by never married woman who reported that they currently or formerly had an intimate partner. In the context of the revised questionnaire module and this report, the term “boyfriend” excludes anyone reported as an intimate partner.

### Measurement of Variables

#### Outcome Variables

The outcome variables for this study were experience of physical violence, experience of emotional violence and experience of sexual violence among women aged 15-49 years currently in an intimate relationship. The outcome variable was measured by self-reported experiences of the women. The operational definition as stated in the original report [9] of three forms of violence is given in Table 1 below.:

**Table 1:** Measure of different types of violence.

| **Forms of Violence** | **Measurements (Questions asked to women if their husband or partner did the following)** |
| --- | --- |
| Physical violence | a. Push, shake, or throw something at her<br>b. Slap her |
|  | c. Twist her arm or pull her hair<br>d. Punch her with fist or something that could hurt<br>e. Kick, drag or beat her<br>f. Try to choke her or burn her on purpose<br>g. Attack her with a knife, gun, or other weapon |
| Emotional violence | h. Said or did something to humiliate her in front of others<br>i. Threatened to hurt or harm her or someone she cared about<br>j. Insulted to make her feel bad about herself |
| Sexual violence | k. Physically force her to have sexual intercourse<br>l. Physically force her to perform any other sexual acts<br>m. Force her with threats or in any other way to perform sexual acts |

Each question was summarized into binary response ‘Yes/No’ to capture the experience of different form of IPV. A value of 1 was given if the event took place (Often, Sometimes, yes but not in the last 12 months) and a value of 0 was given if the act did not take place. The aggregate of a to g was calculated; the woman was considered ‘experiencing physical violence’ if the aggregate was more than 0. If the aggregate was 0 then the woman was considered ‘not experiencing physical violence’. Using similar logic, aggregate of h to j was used for categorizing ‘experiencing emotional violence’ and ‘not experiencing emotional violence’ and aggregate of k to m was used for categorizing ‘experiencing sexual violence’ and ‘not experiencing sexual violence.’

#### Independent Variables

In this study, independent variables included socio-economic variables, partner related characteristics, women empowerment related variables similar to a previous study [13]. The socio-economic variables were age of women, religion, ethnicity, province, region, type of residence, number of family members, wealth index and witnessing parental violence. Partner related characteristics were partner’s education, partner’s occupation, partner’s alcohol consumption, control behavior displayed and the respondent being afraid of their partner. The women empowerment related variables included education, occupation, exposure to the internet, exposure to media, ownership of property, participation in household decisions, attitude towards autonomy of sexual rights and attitude towards justification of beating by partner. [Supplementary file 1].

### Statistical Analysis

We used R version 4.2.0 and RStudio [14] for pre-analytical processing and statistical analysis. We performed weighted analysis to account complex survey design of NDHS 2022 using the survey package [15]. We presented categorical variables as frequency, percentage (%) and 95% confidence interval (CI) whereas numerical variables as mean and standard deviation. We carried out univariate and multivariable logistic regression using stats[14] package to determine factors associated with IPV. The results of the logistic regression were presented as crude odds ratio (COR) and adjusted odds ratio (AOR) and their 95% CI using gtsummary package[16]. We included all the variables with p < 0.2 in univariate logistic regression model to adjust for confounders. No collinearity was found between variables when checked for Variance Inflation Factor for regression models using performance package [17]. A p-value of <0.05 was considered statistically significant.

### Ethical approval

We obtained permission for NDHS 2022 dataset from the DHS program upon registration and providing research title and research purpose. NDHS 2022 received ethical approval for the survey from the Ethical Review Board of Nepal Health Research Council (Reference number: 678, Date: 30th September 2021) and institutional review board of ICF international (Reference number: 180657.0.001.NP.DHS.01, Date: 28th April 2022). In NDHS 2022, written informed consent was obtained from every participant before enrolling them into the study.

## Results

### Characteristics of study population

Table 2 shows the distribution of the respondents by their various socio-demographic characteristics. Of the total 3,853 women, only 0.4% were living together and the remaining 99.6% were married. The age of the respondents ranged between 15 to 49 years with the highest number of respondents in the age group between 35 to 49 years (41.2%). Most of the respondents were from Janajati among prevalent castes, accounting for 36.8%. In terms of provinces, Madhesh (22%), Bagmati (18.4%), Lumbini (17.7%), and Koshi (16.9%) are among the most highly represented provinces. Two thirds of the respondents (66.6%) were from the urban area. The majority of the participants were from the richest wealth quintile (40.8%). The experience of witnessing parental violence in the form of father beating the mother was low with 16.2% of them reporting it.

**Table 2:**
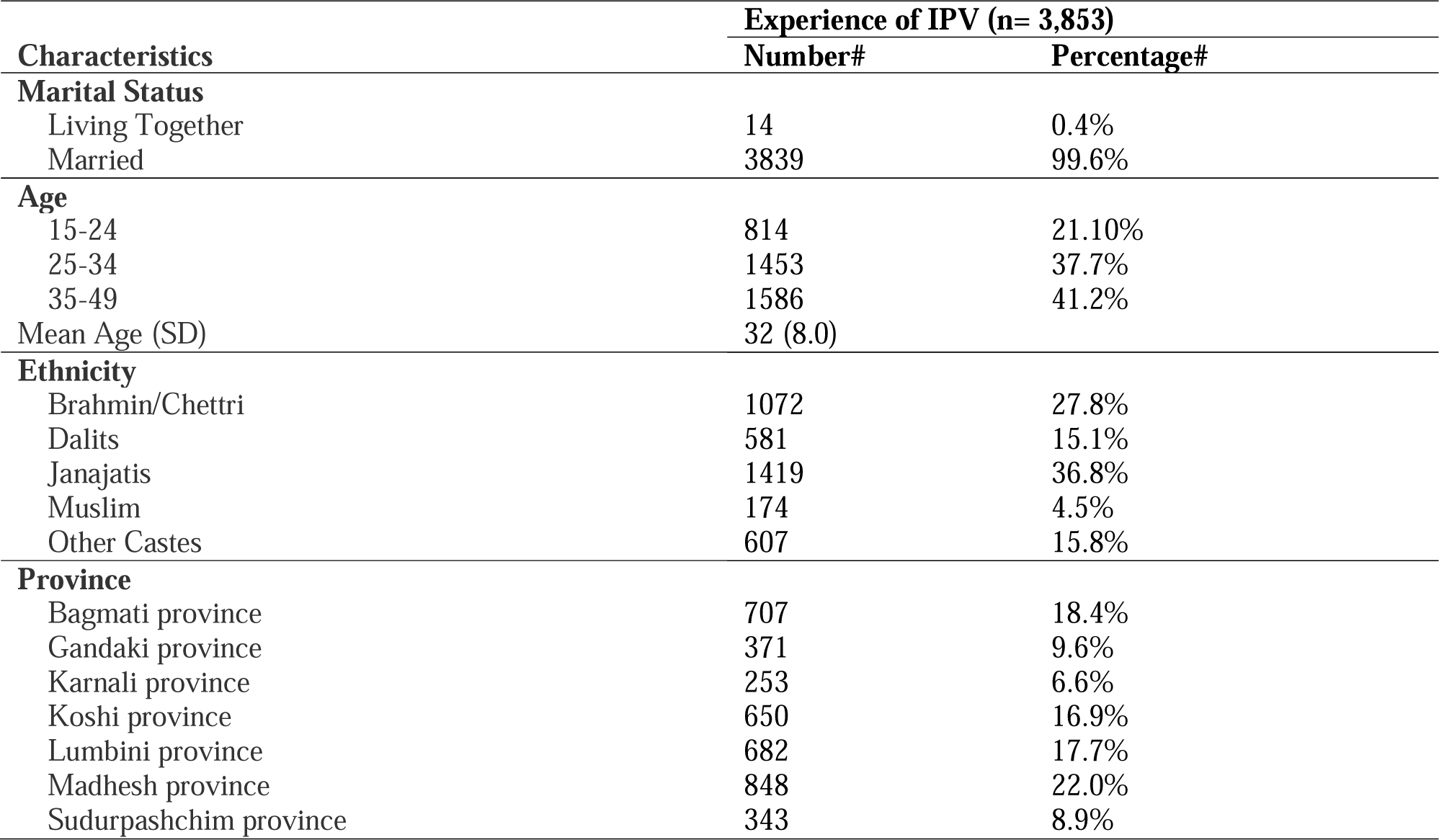
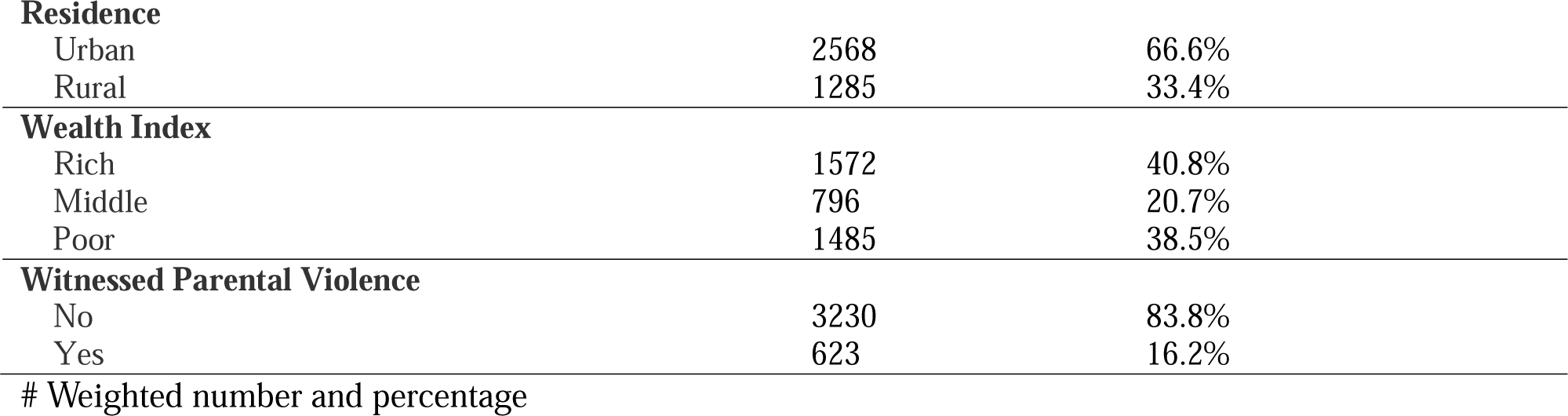
Socio-demographic characteristics of study population.

| Characteristics | Experience of IPV (n= 3,853) |  |
| --- | --- | --- |
|  | Number# | Percentage# |
| **Marital Status** |  |  |
| Living Together | 14 | 0.4% |
| Married | 3839 | 99.6% |
| **Age** |  |  |
| 15-24 | 814 | 21.10% |
| 25-34 | 1453 | 37.7% |
| 35-49 | 1586 | 41.2% |
| Mean Age (SD) | 32 (8.0) |  |
| **Ethnicity** |  |  |
| Brahmin/Chettri | 1072 | 27.8% |
| Dalits | 581 | 15.1% |
| Janajatis | 1419 | 36.8% |
| Muslim | 174 | 4.5% |
| Other Castes | 607 | 15.8% |
| **Province** |  |  |
| Bagmati province | 707 | 18.4% |
| Gandaki province | 371 | 9.6% |
| Karnali province | 253 | 6.6% |
| Koshi province | 650 | 16.9% |
| Lumbini province | 682 | 17.7% |
| Madhesh province | 848 | 22.0% |
| Sudurpashchim province | 343 | 8.9% |
| **Residence** |  |  |
| Urban | 2568 | 66.6% |
| Rural | 1285 | 33.4% |
| **Wealth Index** |  |  |
| Rich | 1572 | 40.8% |
| Middle | 796 | 20.7% |
| Poor | 1485 | 38.5% |
| **Witnessed Parental Violence** |  |  |
| No | 3230 | 83.8% |
| Yes | 623 | 16.2% |

### Husband/ Partner characteristics of the study population

Table 3 shows the characteristics of the respondent’s partner. About 44.2% of the respondent’s partners had received secondary or higher level of education followed by basic level of education (40%) and no education or unknown (15.8%). The majority of the respondent’s partners were engaged in manual work (44.8%) followed by clerical or sales (23.9%), agriculture (18.8%), professional or technical or managerial work (10.2%) and 2.2% did not have any work. Of the total respondents, 64.9% of the respondents reported that their partner did not drink alcohol or were never drunk, and 35.1% reported that they were sometimes drunk or often drunk. About two-thirds of the respondents (66.8%) reported that their partners did not exhibit any controlling behavior and 33.2% had exhibited controlling behavior. Of the total respondents, 43.8% were never afraid of their partner and 56.2% were times afraid of their partner sometimes or most of the time.

**Table 3:** Husband/Partner characteristics of study population.

| **Characteristics** | **Experience of IPV (n= 3,853)** |  |
| --- | --- | --- |
|  | **Number#** | **Percentage#** |
| **Husband/Partner 's Education** |  |  |
| Secondary or Higher | 1703 | 44.2% |
| Basic | 1539 | 40.0% |
| No education | 610 | 15.8% |
| **Husband/Partner 's Occupation** |  |  |
| Professional/technical/managerial | 394 | 10.2% |
| Agriculture | 725 | 18.8% |
| Clerical/Sales/Others | 922 | 23.9% |
| Manual Work | 1727 | 44.8% |
| Not working | 86 | 2.2% |
| **Husband/Partner 's Alcohol consumption** |  |  |
| Does not drink/Never drunk | 2501 | 64.9% |
| Is sometimes/often drunk | 1352 | 35.1% |
| **Control Behavior Displayed by Husband/Partner** |  |  |
| No behavior displayed | 2574 | 66.8% |
| Control behavior displayed | 1279 | 33.2% |
| **Respondent Afraid of Partner** |  |  |
| Never Afraid | 1689 | 43.8% |
| Sometimes or Most of the times afraid | 2164 | 56.2% |

### Empowerment characteristics of the study population

Table 4 shows the empowerment characteristics of woman in the study. The study revealed that most of the respondents (34.7%) had received secondary or higher level of education. There was diverse range of occupational distribution among the respondents with majority (53.6%) engaged in agriculture. A substantial majority of the respondents used the internet (61.6%), were exposed to media (78.2%), and did not own any property (81.5%). The majority (83.7%) of respondents reported participation in household decision making, more than three quarters (77.3%) of respondents reported having the autonomy of sexual rights and 80.6% of respondents agreed that wife beating is not justified.

**Table 4:**
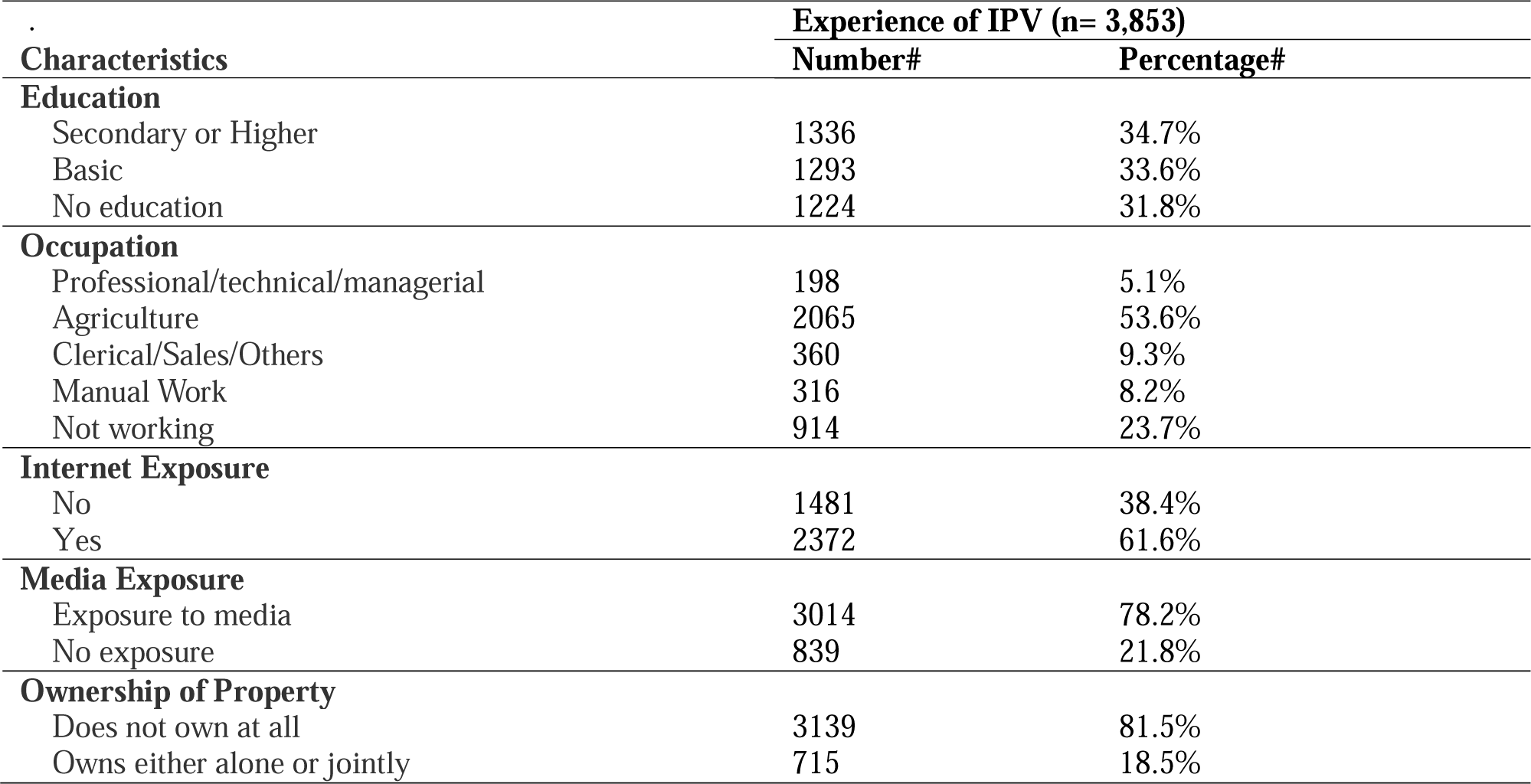
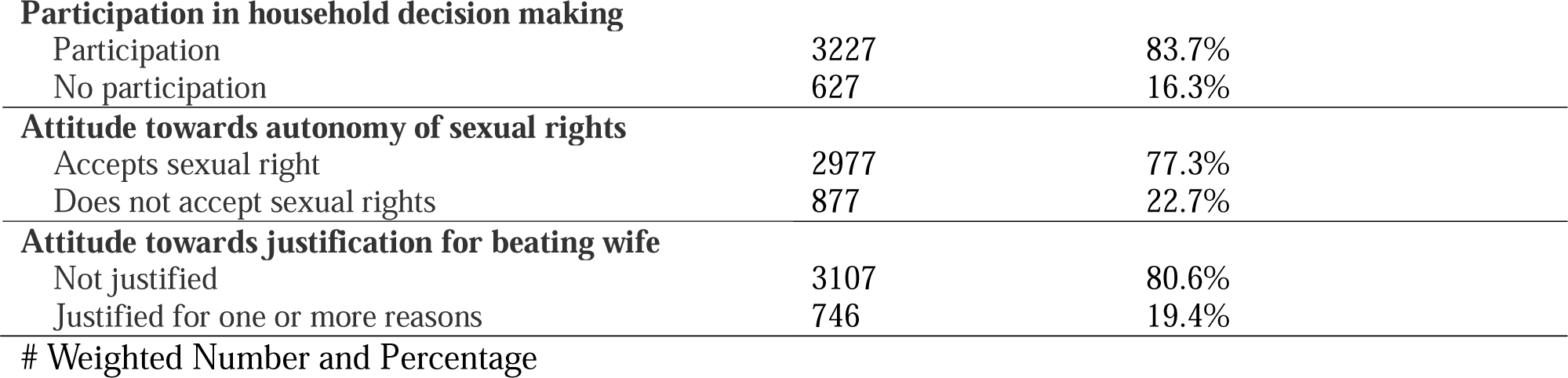
Empowerment characteristics of the study population.

| Characteristics | Experience of IPV (n= 3,853) |  |
| --- | --- | --- |
|  | Number# | Percentage# |
| **Education** |  |  |
| Secondary or Higher | 1336 | 34.7% |
| Basic | 1293 | 33.6% |
| No education | 1224 | 31.8% |
| **Occupation** |  |  |
| Professional/technical/managerial | 198 | 5.1% |
| Agriculture | 2065 | 53.6% |
| Clerical/Sales/Others | 360 | 9.3% |
| Manual Work | 316 | 8.2% |
| Not working | 914 | 23.7% |
| **Internet Exposure** |  |  |
| No | 1481 | 38.4% |
| Yes | 2372 | 61.6% |
| **Media Exposure** |  |  |
| Exposure to media | 3014 | 78.2% |
| No exposure | 839 | 21.8% |
| **Ownership of Property** |  |  |
| Does not own at all | 3139 | 81.5% |
| Owns either alone or jointly | 715 | 18.5% |
| **Participation in household decision making** |  |  |
| Participation | 3227 | 83.7% |
| No participation | 627 | 16.3% |
| **Attitude towards autonomy of sexual rights** |  |  |
| Accepts sexual right | 2977 | 77.3% |
| Does not accept sexual rights | 877 | 22.7% |
| **Attitude towards justification for beating wife** |  |  |
| Not justified | 3107 | 80.6% |
| Justified for one or more reasons | 746 | 19.4% |

### Prevalence of different forms of IPV

Table 5 shows the prevalence of experience of IPV. Overall, 27.2% of the respondents experienced at least one type of violence from their husband or partner, with physical violence being the most common (23.2%). It was followed by emotional violence (12.8%) and sexual violence (7.1%).

**Table 5:** Experience of different forms of IPV.

| Forms of violence | Experience of IPV (n = 3,853) |  |
| --- | --- | --- |
|  | Number # | Percentage # |
| Emotional Violence | 492 | 12.8 |
| Physical Violence | 895 | 23.2 |
| Sexual Violence | 272 | 7.1 |
| Any IPV (Either Emotional, Sexual, or Physical) | 1,047 | 27.2 |

### Bivariate analysis of IPVs by background characteristics

Table 6 shows the experience of physical, emotional, and sexual violence by explanatory variables. The women of age group 35–49 years had high proportions of physical (24.6%) and sexual (7.7%) violence whereas, women aged 15–24 years had high proportions of emotional violence (13%). The proportion of all three violence namely physical, emotional and sexual violence was higher among the respondents: of Muslic ethnic group (49.3%, 27.5% and 16.3%); Madhesh province (39%, 24.9% and 11.9%); rural place of residence (23.4%, 13.2% and 7.4%); poor wealth index (27.3%, 14.5% and 10%); those witnessing parental violence (41.4%, 23.1% and 14.8%); whose husband/ partner had no education (35.8%, 22.8% and 12.6%); whose husband/ partner did not have any work (38.8%, 29.4% and 11.5%); whose husband/ partner was sometimes or often drunk (37.7%%, 21.8% and 13.1%%); whose husband/ partner displayed one or more controlling behavior (44.9%, 30.2% and 17.7%); who were afraid of their husband/partner sometimes or most of the times (32.2%, 18.2% and 11%); who had no education (33.4%, 18.3% and 9.9%); who were involved in manual work (31.7%, 16.5% and 10.4%); who didn’t have exposure to internet (27.2%, 14.3% and 9%); who didn’t have exposure to any media (32.3%, 23.3% and 10.1%); who had no ownership of property (23.9%, 12.9% and 7.1%); who had no participation in household decision making (27.3%, 20.3% and 10%); who didn’t believe in autonomy of sexual rights (35.4%, 21.6% and 12.5%) and respondents who agreed that wife beating is justified for one or more reasons (28.2%, 13.7% and 8.1%). Chi-square analysis indicated that respondents who experienced all forms of physical, emotional and sexual violence had significant association with explanatory variables such as; ethnicity, province, wealth index, witnessing parental violence, husband/ partner’s education, husband/ partners occupation, husband/ partners alcohol use, husband/ partners with controlling behavior, respondents afraid of their husband/ partners, respondents education, exposure to media, respondents participation in household decision making and respondents attitude towards autonomy of sexual rights. The type of women’s occupation was significantly associated with both physical and emotional violence. Women’s exposure to the internet was significantly associated with physical and sexual violence. Whereas women’s attitude towards wife beating was significantly associated with physical violence.

**Table 6:**
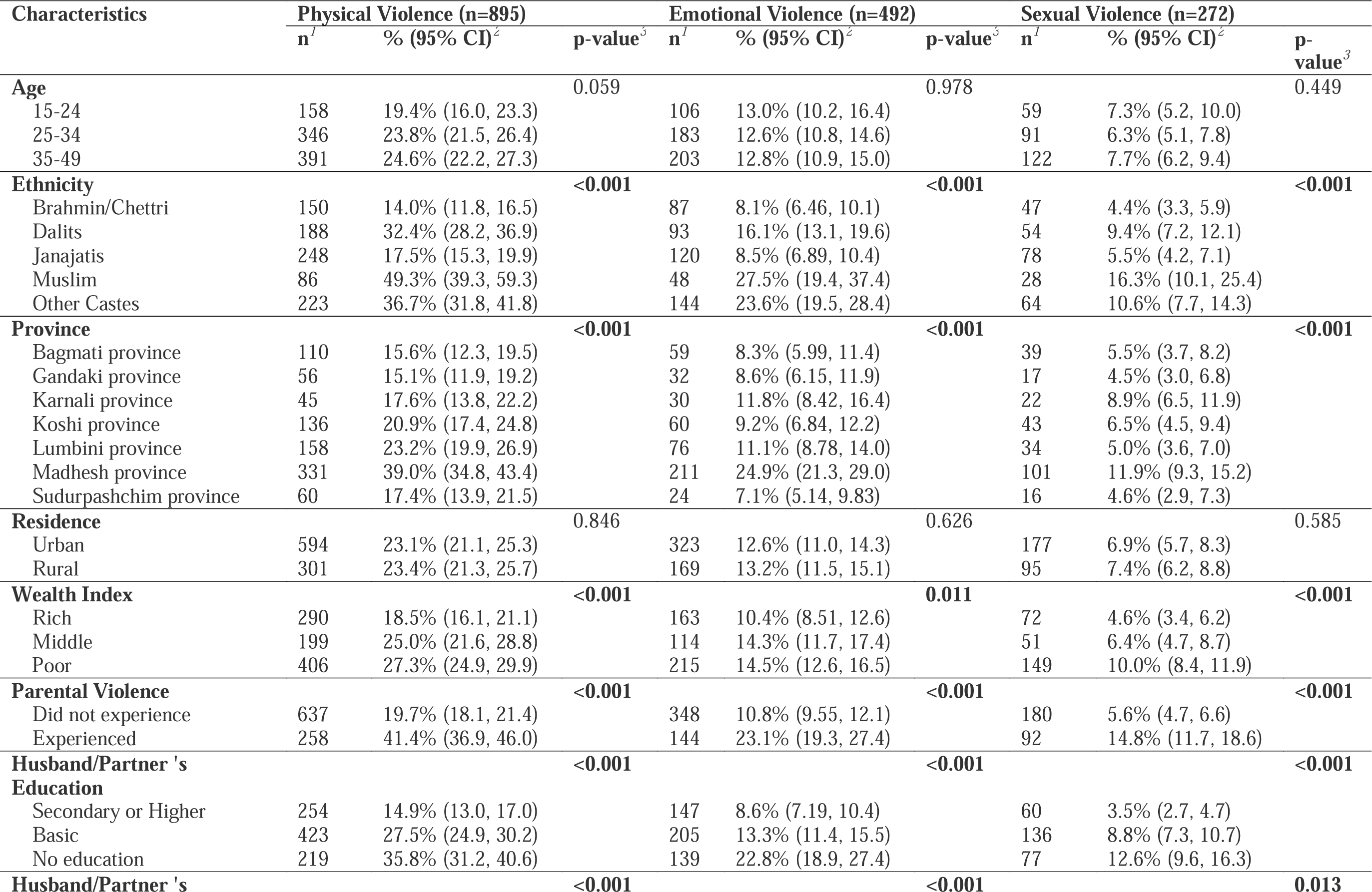
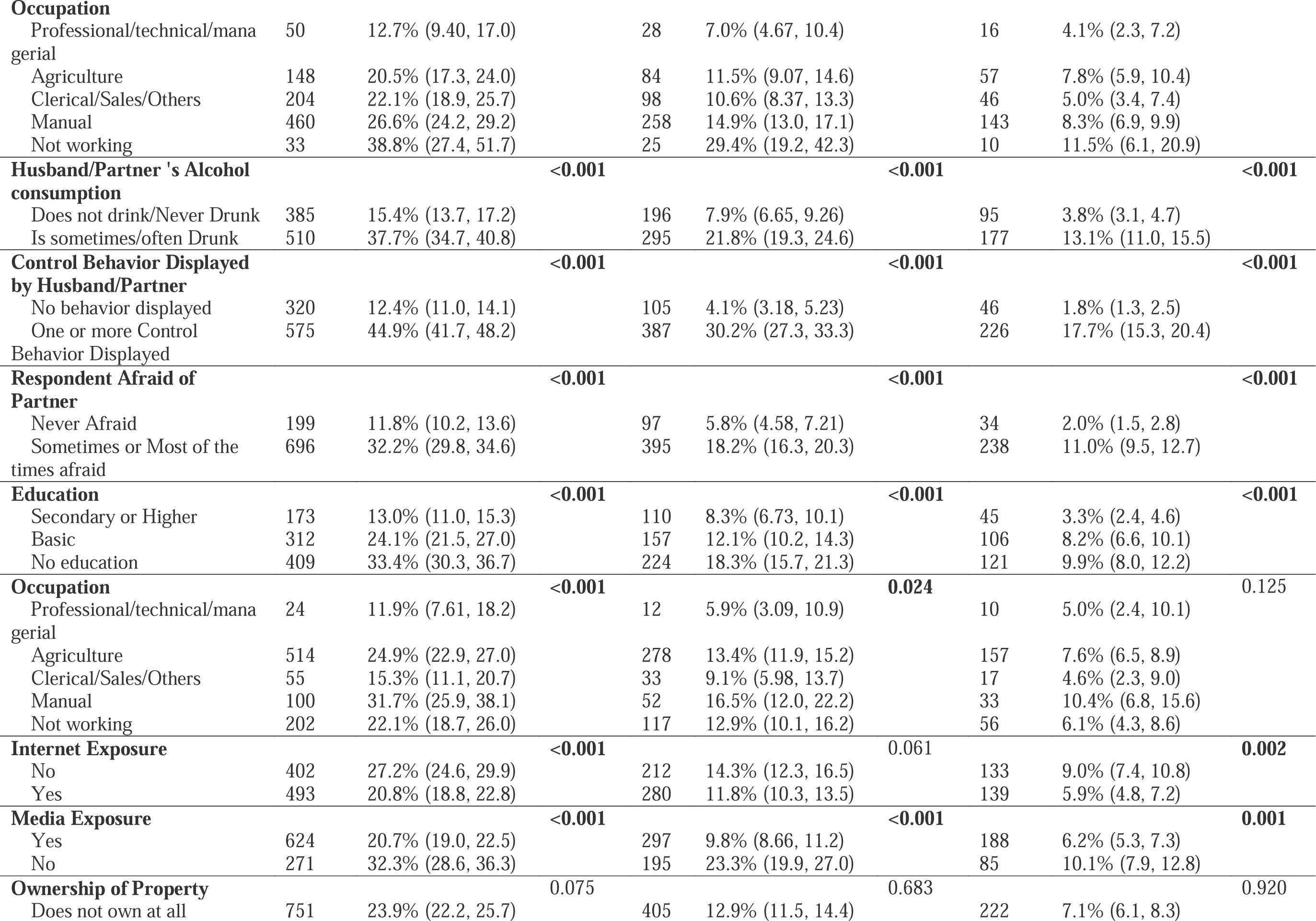
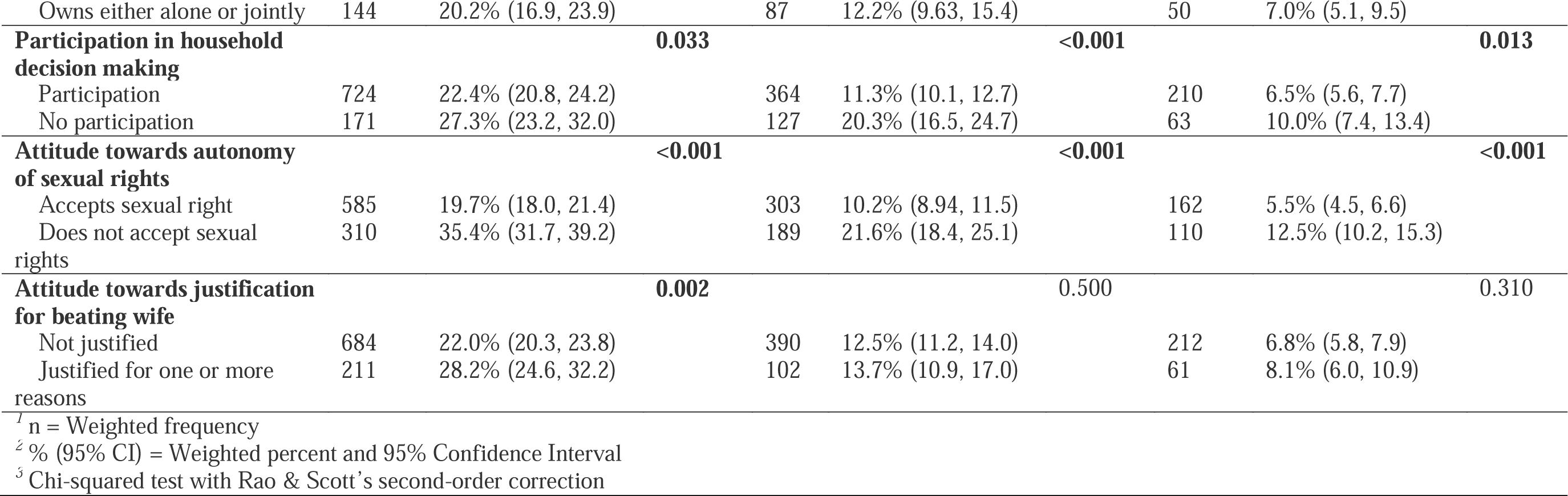
Experience of different IPVs by background characteristics.

### Factors Associated with Experience of different forms of IPV

The logistic regression analysis of the factors associated with experience of different forms of IPVs is shown in Table 7. The regression model showed that age was significantly associated with experience of physical violence with odds being higher with the increase in age i.e., age group 25-34 (AOR: 1.76, CI: 1.37-2.29) and 35-49 (AOR: 2.13, CI: 1.58-2.87). Physical violence was also significantly associated with women’s attitude towards justifications for wife beating with those who justified it for one or more reasons being at higher odds (AOR: 1.23, CI: 1.00-1.52). Similarly, experience of emotional violence was significantly associated with exposure to media; more than two times the odds for those not exposed to any form of media (AOR: 2.34, CI: 1.842.98) compared to those who were exposed to media. Sexual violence was associated with province, with approximately half the odds of experiencing sexual violence in Lumbini province (AOR: 0.47, CI: 0.28–0.77) and Koshi province (AOR:0.58, CI: 0.36-0.93) compared to Bagmati province and women’s husband/partner who were educated up to basic level were more than one and half times more likely (AOR: 1.66, CI: 1.08–2.58) to have experienced sexual violence compared to those whose partners were educated up to secondary or higher level. Also, the likelihood of experiencing sexual violence was almost one and half times higher among those women who did not accept autonomy of sexual rights (AOR: 1.41, CI: 1.05–1.90) than those who accepted their autonomy of sexual rights. Experience of sexual violence was also significantly associated with type of women’s occupation with odds being lower for women not involved in any work (AOR: 0.41, CI: 0.20–0.92) compared to those that were working in professional or managerial or technical fields.

**Table 7:**
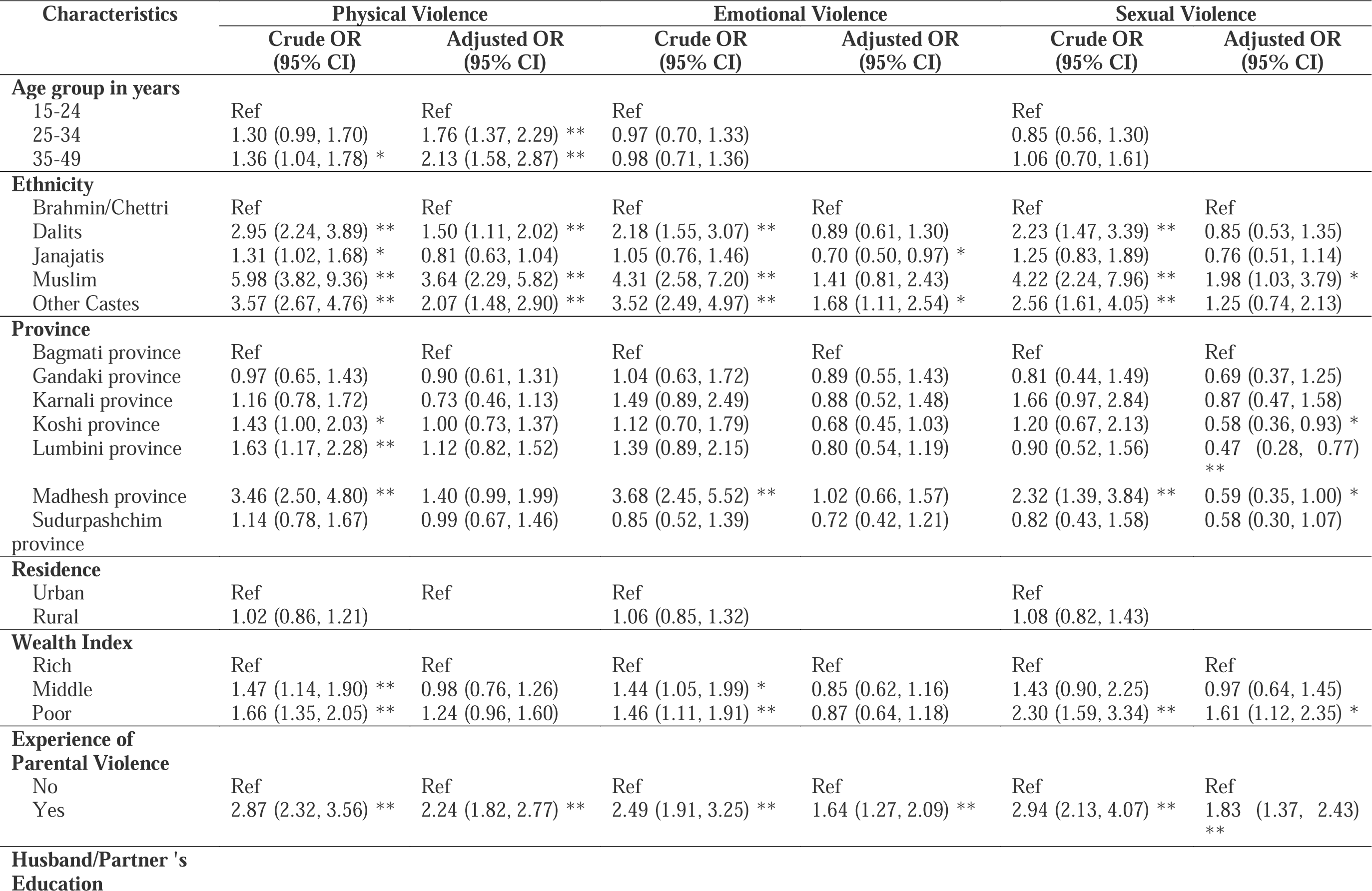
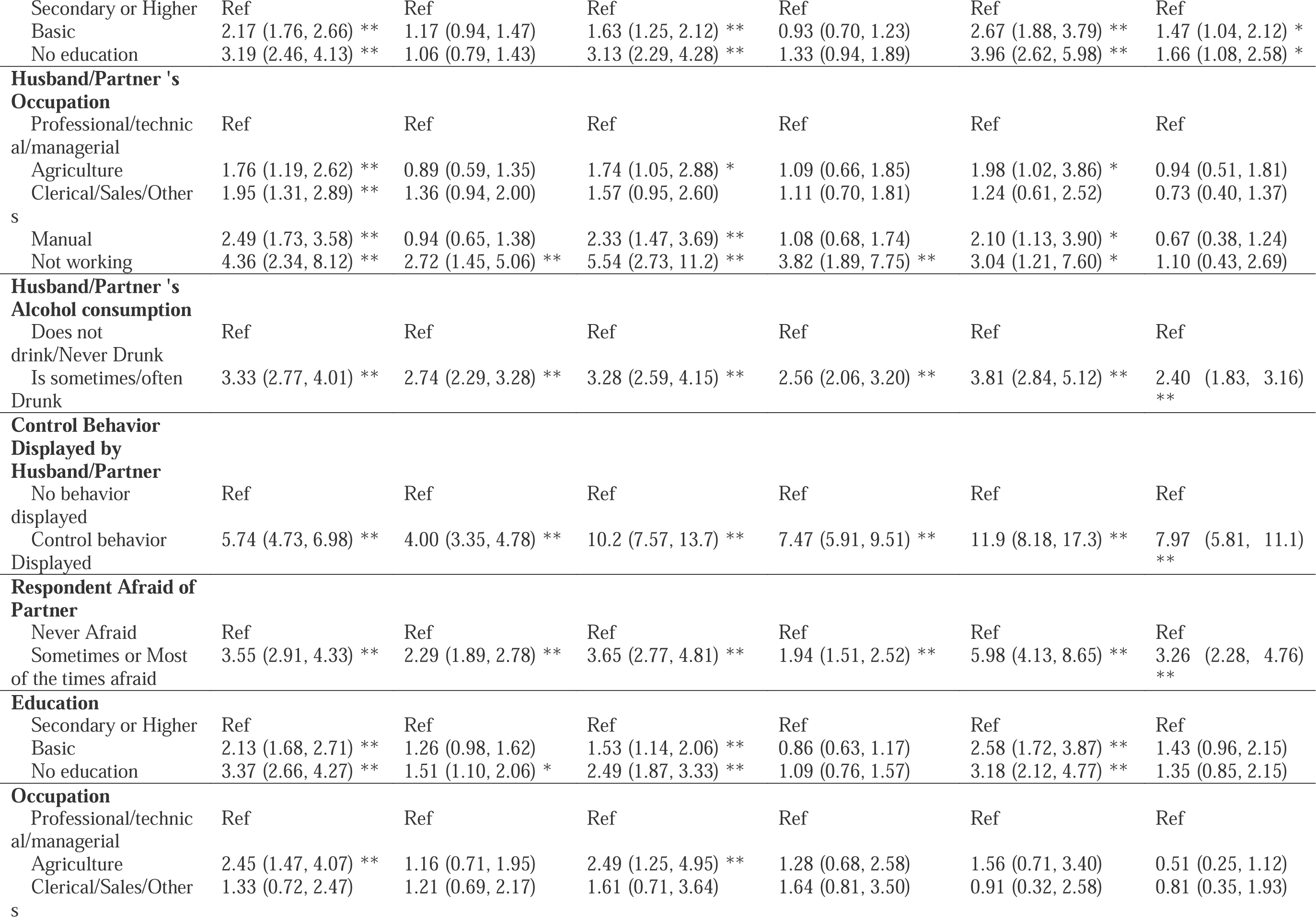
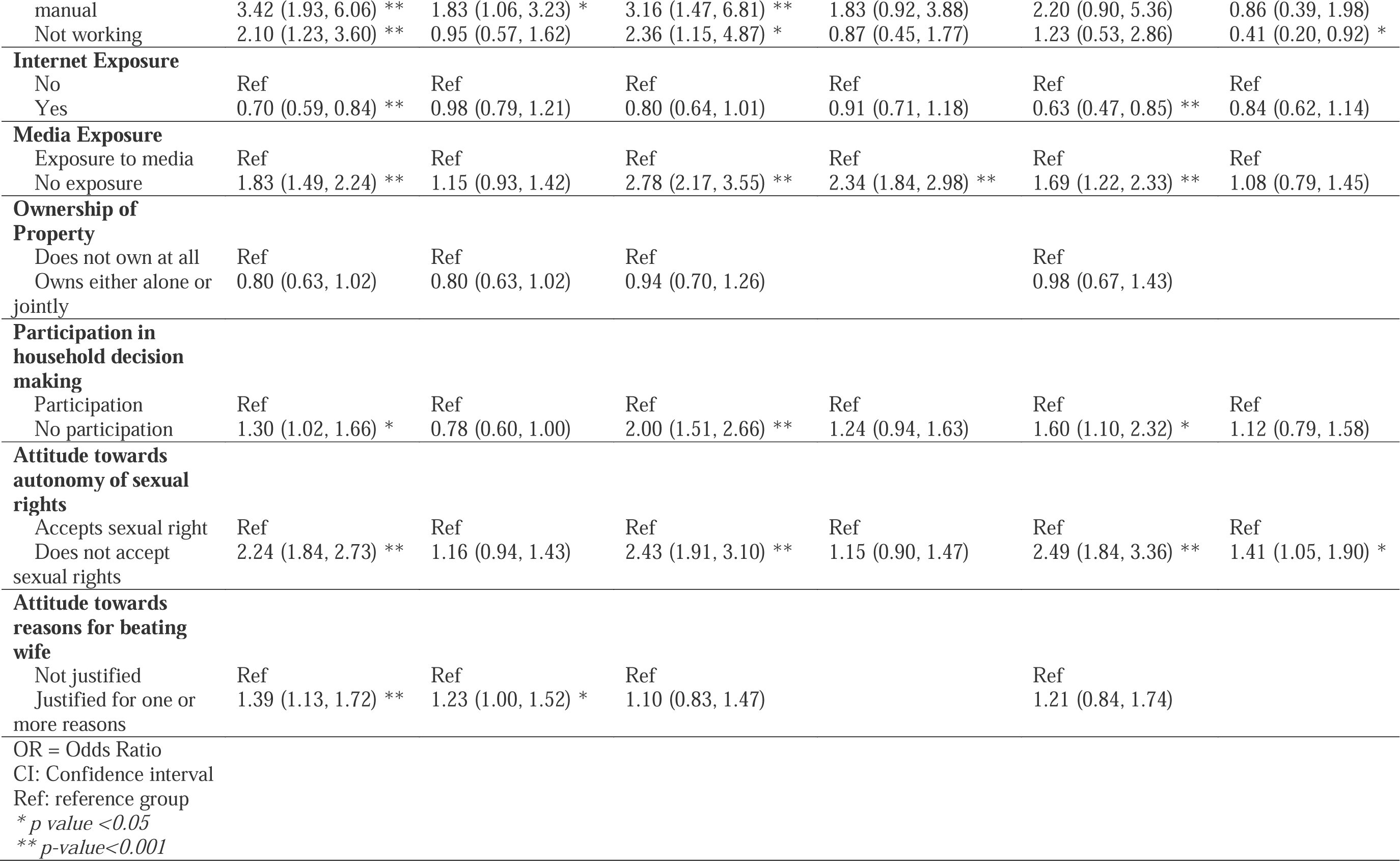
Analysis of factors associated with experience of different forms of IPVs among women in Nepal, 2022 NDHS.

Ethnicity was significantly associated with all forms of IPVs. Women belonging to the Dalit community (AOR: 1.50, CI: 1.11–2.02), Muslim Community (AOR: 3.64, CI: 2.29–5.82) and other caste groups (AOR: 2.07, CI: 1.48–2.90), had significantly higher likelihood of experiencing physical violence compared to women belonging to Brahmins/Chhetri community. Women belonging to the Muslim community (AOR: 1.98, CI: 1.03–3.79) and other caste groups (AOR: 1.68, CI: 1.11–2.54) were also more likely to experience sexual and emotional violence, respectively.

Wealth was significantly associated with sexual violence in the adjusted model with the poor being more likely to experience sexual violence (AOR: 1.61, CI: 1.12–2.35) compared to the rich. Witnessing parental violence was significantly associated with all forms of IPVs with the odds being more than twice (AOR: 2.24, CI: 1.82–2.77), one and half times (AOR: 1.64, CI: 1.27–2.09) and almost twice (AOR: 1.83, CI: 1.37–2.43) more for physical violence, emotional violence and sexual violence respectively compared to those that did not witness parental violence.

Women whose husband/partner did not work were almost three times more likely to experience physical violence (AOR: 2.72, CI: 1.45–5.06) and almost four times more likely to experience emotional violence (AOR: 3.82, CI: 1.89–7.75) compared to those whose husband/partner were working in professional or managerial or technical fields. Similarly, alcohol consumption behavior of partner/husband was also significantly associated with all forms of IPVs. Compared to those whose husband/partner did not consume alcohol or were never drunk, the likelihood of experiencing physical violence was almost three times higher (AOR: 2.74, CI: 2.29–3.28), emotional violence was two and half times higher (AOR: 2.56, CI: 2.06–3.20) and sexual violence was also almost two and half times more (AOR: 2.40, CI: 2.83–3.16) for women whose husband/partner were sometimes or often drunk. Women were also more likely to report IPVs if their husband/partner demonstrated controlling behavior. The odds were almost four times more for physical violence (AOR: 4.00, CI: 3.35–4.78), seven and half times more for emotional violence (AOR: 7.47, CI: 5.91–9.51) and about eight times more for sexual violence (AOR: 7.97, CI: 5.81–11.10) compared to those whose partner did not display any control behavior.

Similarly, women being afraid of their partner was significantly associated with all forms of IPVs. The likelihood of experiencing physical violence (AOR: 2.29, CI: 1.89–2.78), emotional violence (AOR: 1.94, CI: 1.51–2.52) and sexual violence (AOR: 3.se26, CI: 2.28–4.76) was high for those that were sometimes or most of the time afraid of their husband/partner compared to those who were never afraid.

## Discussion

This nationally representative study found that 27% of women in an intimate relationship experience a form of IPV with proportion of women experiencing physical violence, emotional violence, sexual violence being 23%, 12%, and 7%, respectively. The prevalence of various forms of IPV in this study was similar to NDHS 2016 [18] with a notable decrease in emotional and sexual violence as compared to the data from NDHS 2011[19]. Various studies in Nepal report large ranges of IPV prevalence probably due to differences in methodology, sample size and the study area [20–22]. However, the prevalence of IPV could have been under-reported because of societal norms, feelings of shame, embarrassment and the stigma associated with open discourse on marital issues, particularly pertaining to sexual matters [23]. Also, disclosing violence perpetrated by husbands is quite difficult for women because of the culture of silence surrounding men’s acts and the normalization of violence against women [24].

Whilst few studies in low and middle income countries report no association of age with IPV, [25,26] our study found age to be significantly associated with physical violence. Contrary to our finding, the WHO in 2021 reported the highest rate (16%) of IPV occurred among young women aged 15 to 24 [27]. Women belonging to disadvantaged ethnic groups exhibit a higher prevalence of IPV, aligning with findings reported in other studies conducted in Nepal [10,20,28–31].

This study has shown that women belonging to lower socio-economic status families are more likely to experience sexual violence. An analysis from Demographic and Health Surveys in 36 countries also reported higher likelihood of sexual among participants with less household wealth [32]. On the contrary, a study among Iranian women reported significant relationship between socio-economic status and all forms of violence [33]. Similarly, the study showed that women with less educated or unemployed husbands were many times more likely to experience IPV. Low education and unemployment of husbands/ partners have been associated with the experience of IPV in other studies as well [13,34]. Goode’s [35] application of the resource theory by Blood and Wolfe [36] is one of the most cited articles in the literature on why IPV occurs. Goode conceptualizes violence being like a material resource that can be used to gain obedience and compliance in the absence of material resources in the family. Violence or the threat of violence serves as a complement to material resources such as income or education. Thus, this theory leads to the expectation that less educated husbands and husbands with low socio-economic status or income are more likely to perpetrate violence to their partners [37].

Partners who were often drunk or sometimes drunk were significantly associated with all forms of IPV. This finding is supported by a study analyzing demographic and health survey data from 14 sub-Saharan African countries which reported partner’s alcohol use was associated with significant increase in the odds of reporting IPVs in all the countries included in the analysis [38]. Many studies link partner’s alcohol use with perpetration of violence [39–41]. Alcohol consumption is linked with aggressive behavior. A meta-analytic review pooling 22 studies detected a significant overall effect and reported that male participants who consumed alcohol exhibited greater aggressive behavior against females than those who didn’t [42].

The study indicates an association between the attitude towards justification of wife beating with physical violence and the attitude towards the autonomy of sexual rights with sexual violence. Consistent with the application of Bandura’ s social learning theory [43],women internalizing these attitudes are the results of a learned process from observations of social interactions within cultural context where they observe that men are approved of exercising coercion and abuse to instill discipline [44]. Consistent with the findings from Kenya, a husband’s controlling nature was associated with all forms of violence [45]. This reflection also aligns with the attitude towards autonomy of sexual rights as well. IPV was also linked with experience of parental violence similar to another study in Nepal [13]. A study in India showed that women who experienced their father beating their mother were more likely to accept violence [46]. Witnessing violence during childhood may result in normalization or acceptance of violence by women [41]. Childhood experiences of violence at home reinforce normative form of violence for both men and women which subsequently increases the likelihood of perpetration for men and acceptance for women [47]. These findings are consistently reported by various studies [48,49].

Education was a protective factor only for physical violence and the risk of experiencing sexual violence was high for women in professional/managerial/technical occupation. Ownership of property and participation in household decision making was not found to be significant. Our findings contradict the social causation perspective where increasing women’s personal resources such as income and education reduced both recent and longer term probabilities of experiencing any form of IPV [50]. This study’s findings show that empowerment in terms of education and occupation alone cannot guarantee lower risks of IPV. Our finding, however, aligns with the Gender Resource theory which argues that the risk of IPV exists for empowered women as well if the husband is regressive or traditional [37]. The findings call for acknowledgement of marital and societal domains when attempting empowerment related activities against IPV [51]. These findings indicate that a linear relationship doesn’t exist between empowerment characteristics and resources where increased resources would lead to lower risks of IPVs, but the relationship is quite complex [52].

### Strengths and Limitations of the study

The data for this study has been pooled from a national level survey with multi-stage sampling procedure. Sampling weights have been used to adjust for the complex study design so that the data can be made nationally representative. Since this study captures data related to a topic that is still stigmatized in the community, respondents may have been hesitant to report their experiences of IPV, which limited our access to certain details, potentially impacting the depth of our analysis. Also, the WHO’s recommended privacy [12] was ensured to collect the data this. Due to the cross-sectional nature of the study, causal inference cannot be made. Our study also relies solely on quantitative data, and we believe a mix of qualitative and quantitative data is required to help better understand IPV.

## Author Contribution

PMS and ARP were responsible for conceptualization and design of the study. ARP was responsible for acquisition of the dataset. PMS and BA were responsible for data curation and formal analysis. PMS, RP, and GS were responsible for writing the original draft of the manuscript. SG and BL were responsible for writing-review and editing. DJ and SCB were responsible for critically reviewing and supervising the manuscript. PMS is the guarantor of the manuscript. All authors approved the final version of the manuscript.

## Acknowledgement

The authors are grateful to the DHS program for conducting the survey and providing access to the dataset.

## Data availability statement

The data are available publicly in the open-access repository. The data can be downloaded from the official website of ‘The Demographic and Health Surveys’ program. (https://dhsprogram.com/data/dataset/Nepal_Standard-DHS_2022.cfm?flag=0)

