## Supplementary file 1 for "Intimate partner violence in Nepal: Analysis of Nepal Demographic and Health survey 2022"

Table 2: Measurement of independent variables

| **Variables** | **Measurement** |
| --- | --- |
| Age group (in years) | Self-reported age of women at the time of survey, grouped into 15-24 years; 25-34 years; and 35-49 years |
| Ethnicity | Self-reported ethnic affiliation of respondents grouped into Brahmin/Chhetri (Hill Brahmin, Hill Chhetri, Terai Brahmin/Chhetri), Dalits (Hill Dalit, Terai Dalit), Janajatis (Newar, Hill Janajati, Terai Janajati), Muslim (Muslim) and other castes (Other, Other terai caste) |
| Place of residence | Place of residence: Rural, Urban |
| Province | The provincial residence of respondent at the time of survey; Koshi, Madhesh, Bagmati, Gandaki, Lumbini, Karnali and Sudurpaschim |
| Household wealth status | A composite index of household possessions, assets, and amenities, derived using principal component analysis, grouped as Poor (Poorest and Poorer); Middle; and Rich (Richer and Richest) |
| Witnessing parental violence | Self-reported history of witnessing violence in the family measured as, Father ever beat her mother: Yes; No (No, Don’t Know). Not knowing if the father ever beat the mother has been classified as No since, the respondent hasn’t witnessed any parental violence |
| Husband/Partner Education | The highest level of education attained by the respondent’s husband/partner; No education, Primary, Secondary or Higher (Secondary, Higher Education). 49 responses categorized as “Don’t know” have been classified into No education |
| Husband/Partner Occupation | Categorized into not working, professional/technical/managerial, agriculture, clerical/sales/others, and manual (skilled or unskilled); 24 responses categorized as “Don’t know” have been recategorized as others. |
| Husband/Partner Alcohol use | Respondent reporting of partner’s frequency of alcohol consumption, measured as; Doesn't drink/Never Drunk and Is sometimes/often Drunk |
| Women afraid of husband/partner | Self-reported behavior of women being afraid of their husband/partner as; Never afraid; Sometimes or Most of the time afraid |
| Control behavior displayed by husband/partner | A composite variable of self-reported five control behavior displayed by husband/partner (is jealous if she talks to other men, wrongly accuses her of being unfaithful, doesn’t permit her to meet her female friends, tries to limit her contact with her family, insists on knowing where she is at all times). This was grouped into: No behavior displayed, and control behavior displayed. |
| Education of women | The highest level of education attained by respondents: No Education; Primary; Secondary or Higher |
| Occupation of women | Self-reported occupation categorized as into not working, professional/technical/managerial, agriculture, clerical/sales/others, and manual (skilled or unskilled) |
| Exposure to media | A composite variable derived from the frequency of access to newspaper/magazine, radio, and television, grouped as, No exposure, Exposure to media |
| Exposure to internet | Categorized as No and Yes based on frequency of access. Never categorized as No, and Yes, last 12 months; Yes, before 12 months; Yes, can’t establish when grouped as Yes |
| Ownership of property | A composite variable derived from the respondent’s ownership of house, land or both alone or jointly with husband, grouped as: Does not own (Does not own at all); Owns a property (Owns house or land or both either alone or jointly) |
| Women’s participation in household decision making | A composite variable measured from women’s participation (alone or with husband) in making three household decisions (access to healthcare, major household purchases and visit her family or relatives) grouped into No participation; Participation in decision making |
| Attitude towards autonomy of sexual rights | A composite score of women’s abilities to negotiate sexual relations with husband measured from responses of two questions: Women can refuse sex if they don’t want to; and can ask their partner to use a condom. The score ranges from 0 to 2, measured as attitude towards the autonomy of sexual rights. A score of 2 meant they accept autonomy of sexual rights and a score of 0 or 1 meant they don’t believe in sexual rights |
| Attitude towards wife beating (no. of reasons for which wife beating is justified) | A composite variable reflecting women’s attitudes towards beating by their partner for each of the following reasons (Goes out without telling her partner, neglects the children, argues with the partner, refuses to have sexual intercourse with the partner and burns the food), grouped as Not Justified; Justified for 1 or more reasons |
